# Phenotypic Findings Associated with Variation in Elastin

**DOI:** 10.1101/2024.09.10.24313340

**Authors:** Anne Justice, Melissa A. Kelly, Gary Bellus, Joshua D. Green, Raza Zaidi, Taylor Kerrins, Navya Josyula, Teresa R. Luperchio, Beth A. Kozel, Marc S. Williams

## Abstract

Variation in the elastin gene (*ELN*) may contribute to connective tissue disease beyond the known disease associations of Supravalvar Aortic Stenosis and Cutis Laxa. Exome data from MyCode Community Health Initiative participants were analyzed for *ELN* rare variants (mean allele frequency <1%, not currently annotated as benign). Participants with variants of interest underwent phenotyping by dual chart review using a standardized abstraction tool. Additionally, all rare variants that met inclusion criteria were collapsed into an *ELN* gene burden score to perform a Phenome-wide Association Study (PheWAS). Two hundred and ninety-six eligible participants with relevant *ELN* variants were identified from 184,293 MyCode participants. One hundred and three of 254 living participants (41%) met phenotypic criteria, most commonly aortic hypoplasia, arterial dilation, aneurysm, and dissection, and connective tissue abnormalities. *ELN* variation was significantly (P <2.8×10^−5^) associated with “arterial dissection” in the PheWAS and two connective tissue Phecodes approached significance. Variation in *ELN* is associated with connective tissue pathology beyond classic phenotypes.

**eTOC Blurb:** Carriers of variants of interest in the elastin gene (*ELN*) were evaluated for presence of findings that could be associated with the variation. Chart review and Phenome-wide Association Studies were used. Results are consistent with variation in *ELN* being associated with findings affecting elastic tissues beyond classic phenotypes.

## INTRODUCTION

Elastin (*ELN*) encodes a connective tissue protein of the same name (ELN). Secreted as a soluble monomer, it is crosslinked by lysyl oxidase in the extracellular space into a highly insoluble polymer through its lysines^1^ and serves as a component of elastic fibers. Elastic fibers provide recoil to tissues that stretch, including the skin, blood vessels, and lungs. *ELN* has known associations with two well-described genetic conditions.

Haploinsufficiency for *ELN* causes the disease supravalvar aortic stenosis (SVAS, OMIM#185500 https://www.omim.org/entry/185500), characterized by arterial narrowing and stiffness, as well as hypertension. Missense and frameshift changes in the C-terminal part of the gene lead to autosomal dominant cutis laxa (OMIM#123700 https://www.omim.org/entry/123700). Cutis laxa is a multisystem disorder consisting of loose skin, premature emphysema, arterial tortuosity, aortic aneurysm, and inguinal hernia.

Case studies and population-specific genome association studies have linked variation in *ELN* to phenotypes such as chronic obstructive pulmonary disease^2^, aneurysms^3,4^, aortic diameter^5^, and hypertension^6^, suggesting a broader phenotypic spectrum. A recent genome-wide association study (GWAS) studying herniae, identified *ELN* (and other genes in collagen and elastin pathways) as being associated with inguinal hernia^7^. These studies, while helpful in assessing the contribution of *ELN* to disease, have limitations. Family studies generally focus on more distinct or severe phenotypes and, by their nature, select for more highly penetrant variants. GWAS depend on hypotheses about the type of disorder more likely to be associated with genetic variation based on pre-existing knowledge and well-characterized phenotypes, thus are less likely to identify novel phenotypes. GWAS use genotyping arrays that do not include all variation in the gene and use imputation which is less accurate and poorly powered for rare genetic variants likely to be pathogenic.

Large, unselected populations with exome or genome sequence (ES or GS) data tied to electronic health record (EHR) data are becoming more common and are available for research^8^.

This supports identification of most variants in a gene of interest allowing comparison of the full range of phenotypes in variant carriers to those in the population that do not carry any variants. This approach has the potential advantages of reducing selection bias and being more agnostic to what is currently known about phenotypes associated with genetic variation, thus promoting novel gene-phenotype discoveries. The purpose of this study was to apply this genotype-first approach to a large, unselected population with ES data to examine phenotypic associations of variation in *ELN*.

## MATERIALS AND METHODS

### Population Studied

Participants were identified through the Geisinger MyCode® Community Health Initiative Study (MyCode). MyCode is a biobank of samples and linked EHR data from over 300,000 patient-participants who have been recruited throughout the Geisinger healthcare system irrespective of phenotype^9,10^ (“MyCode ScoreCard,” https://www.geisinger.org/precision-health/mycode). Participants consent to broad health-related research including genetic analysis, and a subset of consented participants’ exomes (n=184,293 as of September 1, 2023) have been sequenced as part of the DiscovEHR program^11^.

The PheWAS study included all consented participants of all ages living and deceased, while the detailed chart review was limited to living adult participants defined as aged 18 and older.

### Sequencing

Sample preparation, quality controls, and ES were carried out at the Regeneron Genetics Center as previously described^11,12^. Exome capture was performed using NimbleGen SeqCap EZ VCRome for the first 60,960 participants and Integrated DNA Technologies (IDT) xGen kits for the remainder of the participants according to the manufacturer’s protocol. Multiplexed samples were sequenced using 75×75 basepair (bp) paired-end sequencing on an Illumina v4 HiSeq 2500 or the NovaSeq 6000 platform. All reads were aligned to the GRCh38 reference genome using the Burrows-Wheeler Alignment tool^13^. Variants for were called using weCall v1.1.2 [https://github.com/Genomicsplc/wecall], followed by joint calling and harmonization across freezes using GLnexus^14^. Variants were excluded based on genotype quality (GQ ≤ 20), depth of coverage (DP ≤ 7), and allelic balance (AB ≤ 0.15).

### Variant Identification

Participants in MyCode with ES data were analyzed for sequence variation with a minor allele frequency (MAF) of <1% within *ELN*. Variants were annotated with Ensembl Variant Effect Predictor (VEP v100) and reviewed based on variant location, predicted impact, gnomAD MAF, in silico predictions, ClinVar, and literature reports.

Variants annotated as benign or likely benign in ClinVar, or that resulted in no amino acid change (i.e., synonymous) were excluded from analysis. From those rare non-benign variants we selected variants with the highest potential to impact the protein if they satisfied one or more of the following criteria: 1) protein truncation (e.g., nonsense, frameshift, canonical splice), 2) removal or addition of a Lysine (a residue critical for cross-linking), 3) REVEL score of ≥0.75, or a SpliceAI score of ≥0.2. In addition, we also included variants previously reported in at least one individual with a phenotype consistent with current understanding of *ELN*-associated disease, in ClinVar and/or literature (HGMD v2022.2 and Google scholar Boolean variant-based searches). All candidate variants that met minimum QC thresholds were confirmed by an outside laboratory using an orthogonal technique excepting variants in deceased participants.

Confirmed variants were independently interpreted by the outside laboratory and all were classified as variant of uncertain significance (VUS), except for the single nonsense variant classified as likely pathogenic (LP). There were no discordant interpretations between the outside laboratory and the study variant scientist (MK).

Common variants across the genome were used to infer family relationships using PRIMUS (Pedigree Reconstruction and Identification of Maximally Unrelated Set)^15^ to identify family networks/pedigrees including all 1^st^ degree relatives to inform the number of family members with a shared variant. Additionally, PRIMUS was used to identify a maximally unrelated subset of participants, up to 2^nd^ degree, to include in PheWAS analyses.

### Chart Review

All living participants with variants of interest underwent EHR review by two reviewers using a standardized abstraction tool (Table S1) that encompassed phenotypic findings in relevant organ systems defined by the study team based on published literature and clinical experience. Deceased participants had a limited review to determine if the cause of death was potentially attributable to an elastinopathy (e.g., arterial dissection, aneurysm rupture). Reviewers were blinded to variant type. All discrepancies were reviewed by a clinical geneticist (MSW) and resolved in the case of simple error or omission, or by consensus in the case of discrepant interpretation. Absent validated diagnostic criteria to define disorders associated with pathogenic variation in *ELN*, phenotype positive individuals were subjectively defined as participants with one or more **major** phenotypic findings in one or more systems associated with elastin disorders (**bold** in Figure 1) or two or more non-major phenotypic findings in two or more systems (normal type in Figure 1). Family history available in the EHR was also reviewed and documentation of a family history of major phenotypic features was captured. Presence of a family history of one or more major phenotypic features was scored as a major feature for the participant, while presence of a family history of non-major phenotypic features was scored as a minor feature for the participant.

**Figure 1.**
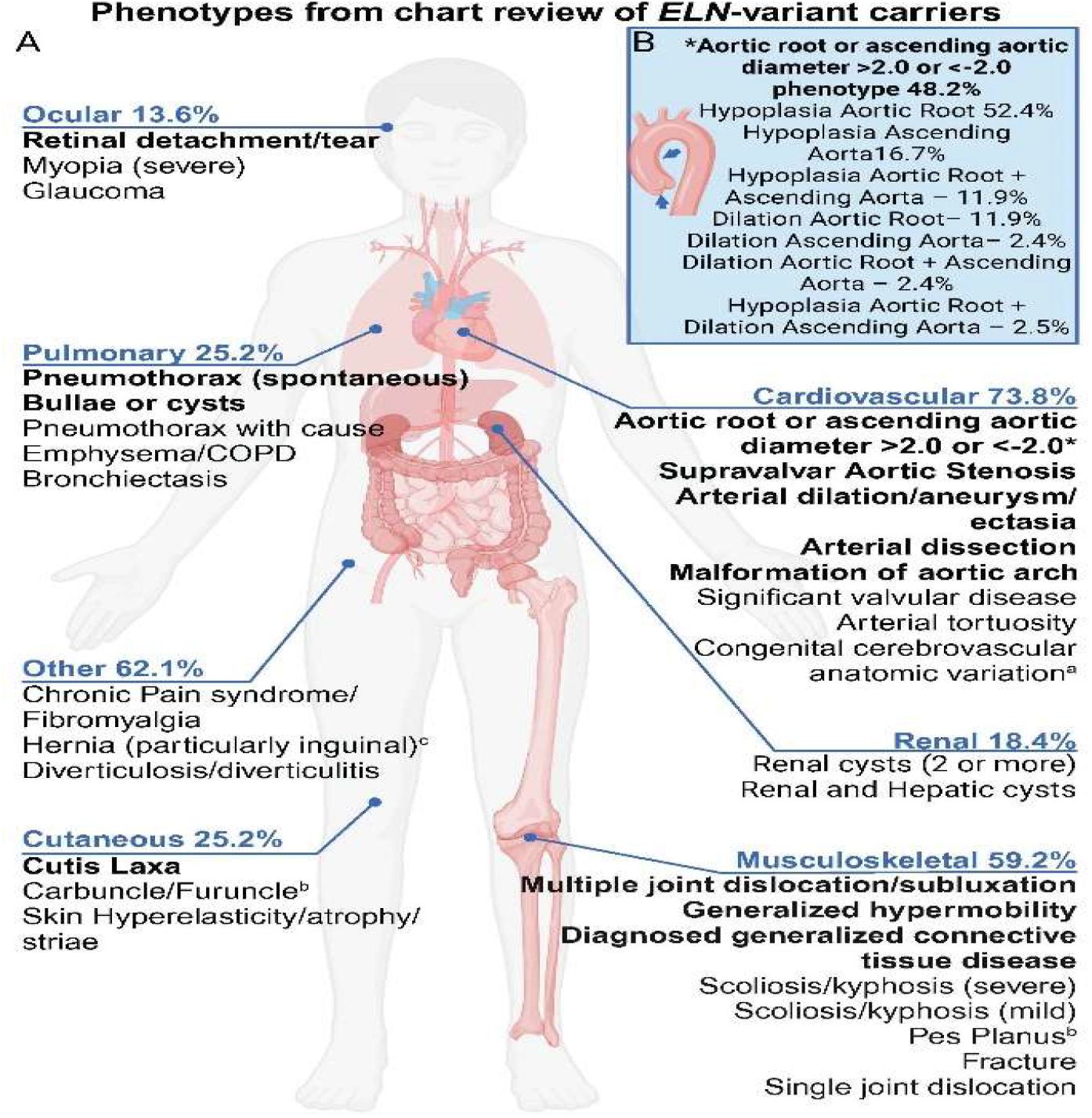
Summary of phenotypic findings from chart review of *ELN* variant carriers. Figure 1A **Bold text** indicates major phenotypic feature. ^a^Not used for phenotype scoring. ^b^Added after initial PheWAS. ^c^Based on GWAS of hernias^7^. Percentages are derived from the number of participants with the specific phenotypic feature divided by all participants with any phenotypic feature (N=103) multiplied by 100. Figure 1B Percentages of participants with an abnormal aortic root and/or ascending aorta. Forty-two participants had an aortic phenotype out of 87 participants with echo measurements to review (48.2%). The percentages of each aortic phenotype is calculated by the number of participants with that phenotype divided by the total number of participants with an aortic phenotype (N=42). Not every echo had measurements for both aortic root and ascending aorta. See Table S3 for additional details. Adapted from “Cancer-Associated Comorbidities”, by BioRender.com (2004). Retrieved from https://app.biorender.com/biorender-templates

#### Aortic Size Normalization

To quantify the aortic diameter measurements in the absence of matched controls, Z-scores were calculated to allow comparison to population norms. Both calculators incorporate the variables of age, biologic sex, height, and weight (to determine body surface area) to standardize measurements. For the aortic root diameter, the Z-score calculator for adults published by the National Marfan Foundation was used (https://marfan.org/dx/z-score-adults/). For the ascending aorta diameter, the Aorta Calculator from the Yale School of Medicine was used (https://medicine.yale.edu/surgery/cardio/research/aorta_calculator_v6_by_tt_370821_4431_v2.xlsx). A Z-score less than -2.0 was classified as hypoplastic and a Z-score of greater than +2.0 was considered dilated. Classification as an aneurysm was based on clinician notes. A Z-score of greater than 2.0 or less than -2.0 for either the aortic root or ascending aorta diameter was considered a major cardiovascular phenotypic feature (Figure 1).

### Phenome-wide Association Study (PheWAS)

To identify novel phenotypes or those not covered in our chart review potentially associated with *ELN* variation, we performed a phenome-wide association (PheWAS) with a simple unweighted burden score for *ELN* variants. For this analysis, we restricted to an unrelated subset of individuals (N up to= 113,513 [195 *ELN* carriers]), including only one member from each family network up to 2^nd^ degree family networks reconstructed using PRIMUS.^15^ Standardized and validated phenotypes (PheCodes) were defined based on the PheCode Map v1.2^16,17^, by extracting all relevant ICD-9-CM and ICD-10-CM codes (Table S2) from all emergency department, inpatient, outpatient, and tele-medicine encounters. PheCode cases were defined by the presence of a minimum of two encounters containing the code and without relevant exclusions coded while PheCode controls exhibited absence of that code and any exclusion criteria. We excluded PheCodes with less than 20 cases and 20 controls from these analyses. A total of 1,781 PheCodes remained following mapping and exclusions. All PheCode mapping, association analyses were carried out using the R package PheWAS v1.0 using logistic regression and adjusting for most recent encounter age, gender (for non-gender specific PheCodes), and the first five principal components using genome-wide common variants to control for population structure. Due to the heterogeneity that may be introduced by conducting PheWAS across race/ethnic groups, we conducted a sensitivity analysis by restricting to white/European American participants.

To detect significant enrichment of associations across phenotypic categories, we conducted a Kolmogorov-Smirnov equality-of-distribution test for the distribution of P-values for each category against all others in our primary results including all participants. We present both one-sided P-values for enrichment for smaller P-values and overall exact combined P-value. Enrichment analyses were carried out in STATA/SE v15.1.

Categories are considered significant with a one-sided P <0.05/17 = 2.9×10^−3^, after correcting for the 17 categories.

## RESULTS

### Chart Review

Two-hundred and ninety-six eligible participants with a rare non-benign variant in *ELN* were identified from 184,293 MyCode participants.

Forty-two were deceased or under age 18 leaving 254 living adult participants. The average age of the participants was 58.6 years (range 23-93). Sex was based on the EHR, with the majority being female—158 females to 97 males (1.63:1 F:M ratio). This is a slightly higher ratio than the MyCode population (1.56:1 F:M). Over 95% of participants self-identified as White, non-Hispanic. On average, phenotype positive individuals (n=103) were older (mean difference = 4.7 years) and more often female than those with *ELN* VUS who were phenotype negative. The male:female ratio in those with *ELN* VUS compared with all *ELN* variant carriers was not significantly different (55 females, 48 males 1.15:1 F:M ratio chi-square = 3.33, P = 0.068). One deceased participant had vertebral artery dissection as the cause of death but none of the other deaths were attributable to known elastin-associated phenotypes. No participant with a diagnosis of SVAS or cutis laxa was identified by chart review, including the participant with the C-terminal stop-gain variant (NM_000501.4:c.2071C>T p.(Gln691Ter)). Of the 254 living participants, 103 (41%) met criteria for presence of a phenotype (Table S3). People with missense variants had a slightly higher positive rate (78 positive of 189 total, 41%) compared to those with a splice variant (25 positive of 65 total, 38%) but the difference is not significant. The variants identified and the associated detailed phenotypes are presented in Table S3. Figure 1 shows the type and frequency of phenotypes identified from the chart review.

Findings involving the vascular system were common affecting 76 of the 103 (73.8%) phenotype-positive participants. Sixty-four of these were a major phenotypic finding (Bold Table S3). Most involved the aortic root and thoracic aorta and included hypoplasia, dilation, or aneurysm, although aneurysm of the pulmonary artery and aneurysm and/or dissection of medium sized arteries were also seen (Figure 1B). Two additional participants had no evidence of vascular disease, but both had a parent that died of a ruptured aortic aneurysm (thought to be thoracic). Hypoplasia of either the aortic root, ascending aorta, or both was seen in 35 of the 103 phenotype-positive participants based on Z-scores <-2 of echocardiogram measurements (Figure 1B), exhibiting a much higher proportions of individuals with abnormal Z-scores than the anticipated 2.5%. This was unexpected in that none of the echocardiogram reports mentioned hypoplasia or small diameter. This is an intriguing finding given the association between *ELN* haploinsufficiency and SVAS. Dilation of the aortic root and/or ascending aorta (defined as a Z-score > +2.0) was seen in ∼7% of participants which is also higher than the anticipated 2.5% although a lower magnitude than hypoplasia. These percentages represent lower bounds, as the denominator was all phenotype positive individuals, while the percentages in Figure 1B represent only those with echocardiographic measurements.

Involvement of the musculoskeletal system was present in 59.2% of phenotype-positive participants (61/103). Only 18 participants had major musculoskeletal phenotypic features (Bold Table S3). One participant had a bone dysplasia, metaphyseal acroscyphodysplasia (OMIM #250215 https://www.omim.org/entry/250215), with an associated atlanto-axial dislocation that required repair. This participant carried a splice variant in *ELN*, NM_000501.4:c.542-1G>T seen in nine participants, three of whom were phenotype positive, but had no evidence of bone dysplasia. One participant had a diagnosis of EDS-hypermobility type and carried the NM_000501.4:c.2134G>A missense variant. Six other participants carried the NM_000501.4:c.2134G>A missense variant, five of whom had no phenotypic findings; the other had tortuosity of the carotids but no connective tissue features. Several participants had generalized connective tissue diagnoses without a specific syndromic or molecular diagnosis. These included fibromyalgia, polyarthralgia, Diffuse Connective Tissue disease, and sacroiliitis rheumatoid arthritis with negative Rheumatoid Factor. The only other major findings seen were three participants with retinal detachment and two with pulmonary blebs/pneumatocele (Bold Table S3). Involvement of other organ systems was less commonly observed (details in Table S3).

No participant had more than one relevant variant detected. In the 254 living adult participants, twenty-five missense and sixteen splice variants were annotated as variants of uncertain significance (VUS) while the single nonsense variant was annotated likely pathogenic. Seventy-five participants had the same missense variant (NM_000501.4:c.659C>T p.(Pro220Leu)) that at the time of submission had conflicting annotation in ClinVar (VUS vs. likely benign). This variant was linked to SVAS in 2000 by Metcalfe et al.^18^, based on an affected mother and son. However, subsequent annotations in ClinVar have not confirmed this. In our cohort, 28 of the 75 participants with this variant had a phenotype, though none had a diagnosis of SVAS. However, Z-score analysis of the 28 phenotype-positive carriers of this variant identified 7 with hypoplasia of the aortic root, 1 with hypoplasia of the ascending aorta and two with hypoplasia of both the root and ascending aorta. PRIMUS for the c.659C>T variant showed that 23 of the 75 were related from nine different family networks (see Methods). No other variant was seen in more than 16 individuals (range 1-16). The most individuals from a single family network was four. Five variants carried by greater than five participants had phenotypic features more than 40% of the time: NM_000501.4:c.2032G>A, 10 of 16 (62.5%); NM_000501.4:c.2171A>T, 6 of 14 (43%); NM_000501.4:c.1150G>A, 8 of 12 (67%); NM_000501.4:c.593C>T, 5 of 11 (45%); NM_000501.4:c.484G>A, 6 of 10 (60%). Details in Table S4.

Of the sixteen variants predicted to impact splicing based on position, the most common variant seen was NM_000501.4:c.1621+1G>A. Only two of 16 participants with this variant had a phenotype of interest. Two other splice variants identified in greater than five participants had phenotypic features 40% or more of the time: NM_000501.4:c.1150+1G>A, 5 of 11 (45%); c.1315+1G>A, 3 of 7 (43%). (Table S4). One splice variant, c.1150+2T>C was observed in four participants all of whom were from the same family and all had phenotypic findings (see Discussion). No other variant was observed to have apparent phenotypic segregation within a family.

### Phenome-wide association study (PheWAS)

169,378 MyCode participants with quality-controlled ES data had available phenomic data that met inclusion criteria, including all 296 *ELN* variant carriers. Of those, we identified 113,513 unrelated participants (92.3% with European genetic similarity and 59.3% women) including 195 *ELN* variant carriers (the other 101 *ELN* variant carriers were removed due to the restriction on relatedness). Of the 1,781 PheCodes analyzed, we identified one phenome-wide significant (P<0.05/1,781 test, or 2.8×10^−5^) PheCode, 442.4 “Arterial dissection” (Table 1, Figure 2). This PheCode includes ICD10 codes under the I77.7 branch “Other arterial dissection” and I67.0 “Dissection of cerebral arteries, nonruptured”. Two additional PheCodes, 727.4 “Ganglion and cyst of synovium, tendon, and bursa” and 727.7 “contracture of the tendon (sheath)”, reached suggestive significance (P<2.8×10^−4^). PheCode 727.4 includes ICD10 codes under the M71.3 branch “Other bursal cyst” and M67.4 branch “Ganglion”. PheCode 727.7 includes M62.4 “Contracture of muscle” and members of the M67.0 branch “Short Achilles tendon” (Table S2). The ten most significant associations are presented in Table 1, which includes additional musculoskeletal and dermatological conditions, e.g., 681.6 “Cellulitis and abscess of foot, toe”, 756.3 “Congenital anomalies of muscle, tendon, fascia, and connective tissue”.

**Table 1.** Top 10 associated PheCodes for PheWAS. This analysis includes all participants. SE=Significant Enrichment. OR=Odds Ratio. P-values that reach Bonferroni-corrected significance are shown in italics. ^a^P-values that reaching suggestive significance.

| PheCode | Beta | SE | OR | P-value | N | N<sub>cases</sub> | N<sub>controls</sub> | PheCode Description | PheCode Category |
| --- | --- | --- | --- | --- | --- | --- | --- | --- | --- |
| 442.4 | 2.473 | 0.513 | 11.856 | *1.46E-06* | 85,410 | 196 | 85,214 | Arterial dissection | circulatory system |
| 727.4 | 1.273 | 0.313 | 3.573 | 4.68E-05<sup>a</sup> | 56,791 | 1,938 | 54,853 | Ganglion and cyst of synovium, tendon, and bursa | musculoskeletal |
| 727.7 | 2.791 | 0.726 | 16.289 | 1.21E-04<sup>a</sup> | 54,932 | 79 | 54,853 | Contracture of tendon (sheath) | musculoskeletal |
| 871.1 | 1.461 | 0.421 | 4.311 | 5.18E-04 | 81,150 | 820 | 80,330 | Open wound of hand except finger(s) | injuries & poisonings |
| 681.6 | 1.433 | 0.423 | 4.193 | 6.96E-04 | 71,433 | 838 | 70,595 | Cellulitis and abscess of foot, toe | dermatologic |
| 362.27 | 1.774 | 0.525 | 5.894 | 7.21E-04 | 90,117 | 449 | 89,668 | Drusen (degenerative) of retina | sense organs |
| 751.22 | 1.696 | 0.508 | 5.452 | 8.45E-04 | 103,916 | 428 | 103,488 | Other specified congenital anomalies of kidney | congenital anomalies |
| 613.7 | 1.527 | 0.460 | 4.603 | 9.04E-04 | 101,363 | 661 | 100,702 | Other signs and symptoms in breast | genitourinary |
| 010 | 2.312 | 0.719 | 10.090 | 1.30E-03 | 90,868 | 132 | 90,736 | Tuberculosis | infectious diseases |
| 756.3 | 2.301 | 0.720 | 9.984 | 1.40E-03 | 102,697 | 136 | 102,561 | Congenital anomalies of muscle, tendon, fascia, and connective tissue | congenital anomalies |

**Figure 2.**
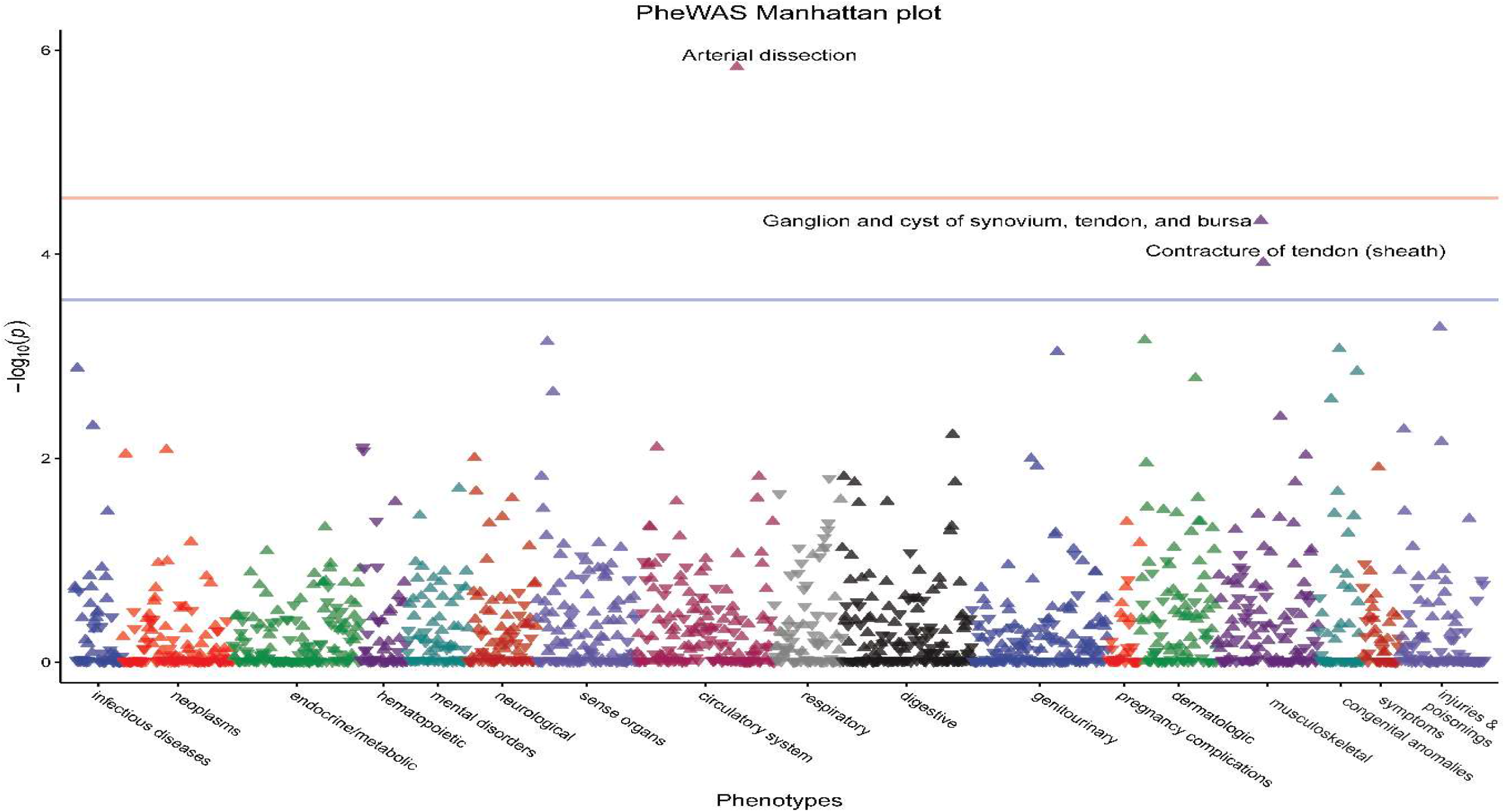
PheWAS Manhattan plot for *ELN* variant burden analysis. Arrows indicate direction of effect with respect to carriers. The red line indicates the phenome-wie threshold of 2.8×10^−5^ and the blue line indicates suggestive significance of 2.8×10^−4^.

While only one dermatologic trait was among our top 10 associations in our enrichment analyses, we identified a significant enrichment for smaller P-values for PheCodes related to dermatologic traits (Table 2 and Figure 3. A total of 11 of the 94 PheCodes in the dermatologic category were nominally associated with *ELN* variation (data not shown). The majority of these PheCodes relate to disorders and diseases of the sweat or sebaceous glands (e.g., 705.8 “Hyperhidrosis”, 706.2 “Sebaceous cyst”) or skin infections (e.g., 686.1 “Carbuncle and furuncle”).

**Table 2.** PheCode category enrichment analysis. Italicized P-values reach Bonferroni-corrected significance level.

| Category | #<br>PheCodes | Smaller |  | 2-sided |  | PheCode with Smallest P-value in Category |  |  |  |  |
| --- | --- | --- | --- | --- | --- | --- | --- | --- | --- | --- |
|  |  | D | P | D | P | PheCode | Description | OR | P |  |
| Dermatologic | 94 | -0.2124 | 3.29E-04 | 0.2124 | 5.34E-04 | 681.6 | Cellulitis and abscess of foot, toe | 4.193 | 6.96E-04 | 3.483147 |
| Mental Disorders | 75 | -0.1655 | 0.02 | 0.1655 | 0.035 | 313.2 | Tics and stuttering | 5.624 | 1.96E-02 | 1.69897 |
| Digestive | 159 | -0.1103 | 0.03 | 0.1103 | 0.055 | 574.1 | Cholelithiasis | 2.164 | 5.82E-03 | 1.522879 |
| Musculoskeletal | 124 | -0.1147 | 0.048 | 0.1147 | 0.089 | 727.4 | Ganglion and cyst of synovium, tendon, and bursa | 3.573 | 4.68E-05 | 1.318759 |
| Circulatory System | 168 | -0.0903 | 0.084 | 0.0903 | 0.158 | 442.4 | Arterial dissection | 11.856 | 1.46E-06 | 1.075721 |
| Neurological | 83 | -0.1216 | 0.097 | 0.1216 | 0.177 | 327.1 | Hypersomnia | 2.136 | 9.89E-03 | 1.013228 |
| Sense Organs | 123 | -0.0937 | 0.135 | 0.0937 | 0.252 | 362.27 | Drusen (degenerative) of retina | 5.894 | 7.21E-04 | 0.869666 |
| Respiratory | 83 | -0.0853 | 0.317 | 0.0853 | 0.583 | 512.9 | Other dyspnea | 0.530 | 1.56E-02 | 0.498941 |
| Congenital Anomalies | 54 | -0.0859 | 0.462 | 0.2367 | 0.005 | 751.22 | Other specified congenital anomalies of kidney | 5.452 | 8.45E-04 | 0.335358 |
| Symptoms | 45 | -0.0688 | 0.66 | 0.1181 | 0.536 | 781 | Symptoms involving nervous and musculoskeletal systems | 2.085 | 1.22E-02 | 0.180456 |
| Neoplasms | 135 | -0.0365 | 0.718 | 0.1952 | 1.25E-04 | 187 | Cancer of other male genital organs | 6.727 | 8.26E-03 | 0.143876 |
| Genitourinary | 167 | -0.0322 | 0.732 | 0.0996 | 0.093 | 613.7 | Other signs and symptoms in breast | 4.603 | 9.04E-04 | 0.135489 |
| Infectious Diseases | 59 | -0.0438 | 0.803 | 0.104 | 0.533 | 010 | Tuberculosis | 10.090 | 1.30E-03 | 0.095284 |
| Hematopoietic | 57 | -0.0354 | 0.871 | 0.1118 | 0.462 | 280 | Iron deficiency anemias | 0.354 | 7.77E-03 | 0.059982 |
| Injuries & Poisonings | 104 | -0.0196 | 0.928 | 0.1388 | 0.042 | 871.1 | Open wound of hand except finger(s) | 4.311 | 5.18E-04 | 0.032452 |
| Endocrine/Metabolic | 156 | -0.0131 | 0.953 | 0.0737 | 0.403 | 272.12 | Hyperglyceridemia | 2.103 | 4.72E-02 | 0.020907 |
| Pregnancy Complications | 39 | -0.0042 | 0.999 | 0.1973 | 0.089 | 649.1 | Diabetes or abnormal glucose tolerance complicating pregnancy | 2.290 | 4.19E-02 | 0.000435 |

**Figure 3.**
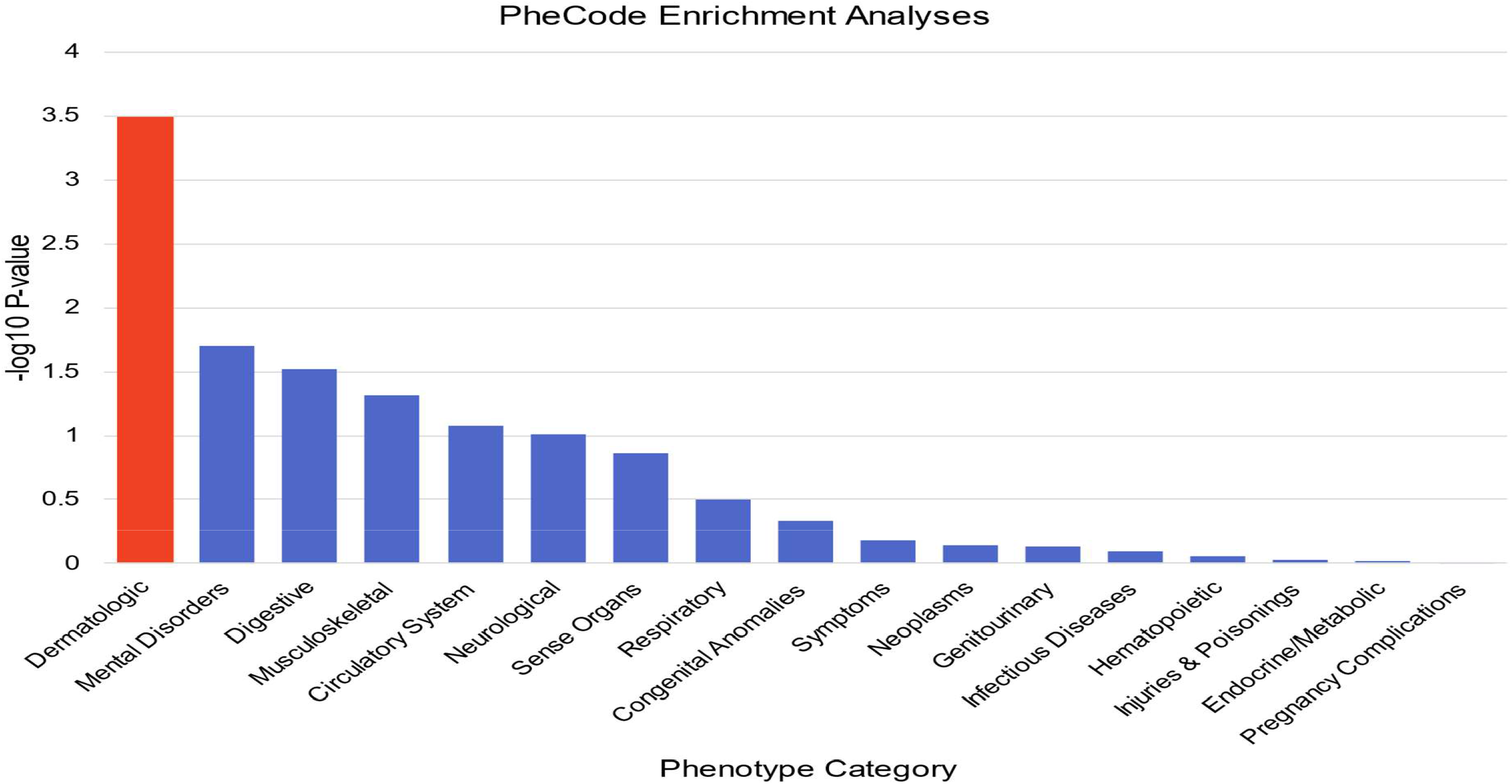
PheCode enrichment analyses. Orange indicates Bonferroni-corrected significance for enrichment of lower P-values compared to all other categories.

As a sensitivity analysis, we conducted a PheWAS restricted to non-Hispanic White / European American participants (Figure S1, Table S5). Our top two associated PheCodes from our primary analysis, 442.4 “arterial dissection” and 727.4 “ganglion and cyst of synovium, tendon, and bursa”, reached the phenome wide significance threshold (P<2.8×10^−5^). No additional PheCodes reached suggestive significance; however, nine of the top 10 PheCodes from our primary analysis were among the top ten associations in our sensitivity analysis, but the suggestive association with 727.7 “Contracture of the tendon (sheath)” was significantly attenuated. Instead, 735.1 “Flat foot” was among the top ten associations in our sensitivity analysis.

## DISCUSSION

This study reports findings from over 250 research participants with variants in *ELN* not selected on the basis of clinical features. The use of chart review and PheWAS allowed study of previously observed phenotypes and promoted identification of novel phenotypes.

### Phenotypic Insights

No cases of SVAS or cutis laxa were identified in the variant carriers. The absence of cutis laxa is not unexpected given that this is usually associated with missense and frameshift variants in the C-terminal part of the gene. The most frequent missense variant studied in this project (NM_000501.4:c.659C>T) is not located in the C-terminal region. The lack of SVAS in the study cohort, while unexpected is likely due to the genetic mechanism. SVAS is caused by haploinsufficiency for *ELN*. The single nucleotide variants identified are not predicted to result in haploinsufficiency, except for NM_000501.4:c.2071C>T which introduces a stop codon at amino acid position 691. This variant is located in exon 32 which is frequently spliced out of the mRNA through a cassette-type alternative splicing, and this could attenuate the functional impact. This participant is a woman in the 26-30-year age range with 18 years of EHR data. She has atopic dermatitis but no other skin findings. She has not had cardiac imaging, so SVAS cannot be ruled out. This finding is corroborated by the SVAS sequencing study (Metcalfe, et al.^18^) that identified only 4 missense mutations in their cohort. No definitive assertion of causality was made by the authors and the only additional criterion applied was absence in 200 controls. Splicing variants are frequently associated with frameshifts, leading to nonsense mediated decay and haploinsufficiency but that may not be true with *ELN* due to the in-frame nature of the exons. While SVAS was not identified, hypoplasia of the aortic root and/or ascending aorta was identified in many participants. This has been previously reported in Williams syndrome and *ELN* haploinsufficiency^19^. Prior studies have not used Z-scores to assess aortic diameter and the focus was either on SVAS or dilation/aneurysm. Based on the echocardiogram reports it appears that hypoplasia is underreported, as none of the individuals with a Z-score less than -2.0 had hypoplasia noted in the report. Z-scores are used in pediatric echo cardiography, but are not included in adult reports, at least in our institution. If this is the case more generally, aortic hypoplasia may be systematically underreported. The health consequences of milder forms of thoracic aortic hypoplasia have not been studied. Additional research examining the impact of these variants on aortic size and the protein product are needed to understand the potential phenotypic consequences.

Vascular findings were the most common major phenotype identified. Beyond the aortic findings, dilation of the pulmonary artery, aneurysm and dissection of medium-sized arteries, arterial tortuosity, and valvar insufficiency were seen (Figure 1). ELN accounts for more than two-thirds of the dry weight of the aorta^20^, and is present in other arteries, so detailed review of arterial phenotypes was defined in the abstraction guide, raising the potential for confirmation bias. The PheWAS identifying arterial dissection as significant mitigates this concern for this specific phenotype. Additionally, the population prevalence of arterial dissection aggregated across anatomic sites is estimated to be 2.6-2.9 per 100,000 per year. This means that in the MyCode population of 184,293 that was available for study, assuming 10 years of medical data for each participant 48-53 dissection events would be anticipated. Thus, in the 296 variant carriers we would have expected 0.08-0.085 cases if the distribution was random. Many more were seen confirming the PheWAS result. These findings provide further evidence of an essential role for the elastin protein in blood vessel integrity.

Musculoskeletal and connective tissue findings were also common, although only one case of generalized joint laxity clinically diagnosed as EDS Hypermobility type was identified. One participant had a bone dysplasia, clinically diagnosed as metaphyseal acroscyphodysplasia. The genetic etiology of this dysplasia is unknown, but presence of the *ELN* variant is not thought to be causal. The same is true for the participant with a clinical diagnosis of a form of EDS. However, musculoskeletal phenotypes were among the top associated PheCodes in our PheWAS, including 727.4 “Ganglion and cyst of synovium, tendon, and bursa” that was suggestively significant in our primary analysis and phenome-wide significant in our sensitivity analysis.

### Variant specific insights

Variants of uncertain significance create dilemmas for clinicians performing genetic testing for a clinical indication. The ambiguity of the result leads to uncertainty in clinical management creating a decisional burden for the clinician and patient. Resources such as ClinVar that attempt to aggregate and reconcile variant annotation coupled with variant interpretation standards promulgated by professional organizations can help but are still hindered by reliance on results obtained from clinical testing that may have incomplete phenotype information and have an inherent ascertainment bias. Population studies may provide additional insights for both gene-disease associations and variant-specific annotation may resolve some of the uncertainty given larger numbers of variant carriers and less biased ascertainment as illustrated by the following examples.

The study has expanded understanding of the c.659C>T missense variant. This variant has a relatively high frequency in the population (MAF=0.000238). This, coupled with the absence of SVAS in the MyCode population means that the reported segregation of this variant with SVAS (Metcalfe, et al.^18^), may be a coincidental finding. However, roughly one third of carriers of this variant had phenotypic features potentially consistent with an elastinopathy (Table S3). Of those, 10 of 28 (36%) had hypoplasia of the aortic root, ascending aorta or both. Other variant carriers had aneurysm of the distal or abdominal aorta, aneurysms of smaller vessels, and arterial dissection (Table S3), so this variant cannot be dismissed as benign. Case-control studies treating aortic diameter as a quantitative trait coupled with longitudinal measurement of changes in aortic diameter could shed additional light on this variant’s association with aortic size.

Position NM_000501.4:c.1150 may have particular importance. A missense variant (NM_000501.4: p.Gly384Arg) and two splice variants (NM_000501.4:c.1150+1G>A and NM_000501.4:c.1150+2T>C) were seen recurrently and more than 40% of carriers of each variant had phenotypic features (Details Table 3). None of these three variants were reported by Metcalfe et al.^18^, in their series of SVAS patients. Overwater et al.^21^, in a series of 810 people with suspected heritable thoracic aortic aneurysms and aortic dissections (TAAD), while not identifying any *ELN* variants they annotated as P or LP, did identify one patient with the c.1150+1G>A variant and another with the c.1150G>A missense variant (Overwater et al., supplemental table 1).

**Table 3.** Phenotypes associated with variation at position c.1150 in *ELN*.

| Variant | Family Hx | Cardiovascular | Renal | Skin | Lung | Ocular | Connective Tissue | Other |
| --- | --- | --- | --- | --- | --- | --- | --- | --- |
| **NM_000501.4:c.1150G>A (p.GLy384Arg)** |  | Normal aortic root and ascending aorta | Bilateral Renal Cysts | Carbuncle requiring intervention |  |  |  | L inguinal hernia. Multiple pancreatic cysts. Multiple lung granulomas |
|  |  | **Stent distal abdominal aorta and iliac arteries** |  | Carbuncle requiring intervention | **COPD. Multiple blebs** |  | Sacroiliitis. Small hiatal hernia |  |
|  |  | Normal aortic root and ascending aorta. Mitral valve prolapse |  |  |  | **Retinal detachment** | Slight dextro-scoliosis | R inguinal hernia. Diverticulosis |
|  |  | **Dilated aortic root (Z=2.2) Normal ascending aorta.** |  |  |  |  | Baker's cyst R knee | Cystocele Uterine prolapse |
|  |  | **Hypoplastic aortic root (Z=-2.33) Normal ascending aorta.** | Multiple cysts R kidney |  |  |  |  | Umbilical hernia |
|  |  | **Normal aortic root and ascending aorta. MRI mild dilation of root Proximal ascending 45.41 mm sinotubular junction 36 x 33 mm. If used the 45.41 for the ascending aorta Z-score is 3.15 classified as ectasia** |  |  |  |  |  | Diverticulosis |
|  | Two children with arthritis | **Normal aortic root. Abdominal aorta ectasia and tortuosity. Short segment dissection infrarenal aorta associated with mural thrombus** |  |  |  |  | Sacroiliitis | Diverticulosis<br>Small hiatal hernia |
|  |  | **Normal aortic root. Hypoplasia ascending aorta (Z=-3.11). Mild to moderate mitral regurgitation** |  |  |  | Glaucoma | Syringomyelia<br>spinal stenosis grade 1<br>anterolithesis<br>synovial cyst.<br>Bilateral pes planus | Complex regional pain syndrome |
| **NM_000501.4:**<br>**c.1150+1G>A** |  | Normal Aortic root and ascending aorta. Mild aortic regurgitation |  |  | COPD |  | Mild scoliosis thoracic spine | Severe diverticulosis |
|  |  | **Normal aortic root and ascending aorta by measurement. Aortic dilation on report. moderate aortic regurgitation** |  |  |  |  | Subluxation biceps tendon |  |
|  |  | **Hypoplastic aortic root (Z=-3.28)**<br>**Vertebrobasilar dolichoectasia, mild aortic stenosis and regurgitation** |  |  | Restrictive lung disease |  | **Multiple joint osteo-arthritis** | Inguinal and umbilical hernias |
|  |  | **Normal aortic root and ascending aorta. Coronary artery dissection. Mild aortic, mitral, and pulmonary regurgitation. Moderately severe tricuspid regurgitation** |  |  |  |  |  | Severe diverticulosis |
|  |  |  | Multiple bilateral renal cysts |  |  |  | R convex scoliosis, Intervertebral disk prolapse. Shoulder subluxation |  |
| **NM_000501.4:**<br>**c.1150+2T>C** |  | **Thoracic Aortic Aneurysm, Aortic Root replacement due to aortic root abscess and endocarditis, aortic valve stenosis and insufficiency (Rheumatic)** |  |  |  |  | Mild levoscoliosis and spondylosis thoracic spine |  |
|  | Parent mitral valve disease | Normal Aortic root and ascending aorta. Mild mitral regurgitation |  |  | Pneumo-thorax |  |  | Chronic pain syndrome |
|  | Parent Ruptured diverticulum | Normal Aortic root and ascending aorta |  |  |  | Chronic Narrow Angle Glaucoma | Cervical Spondylosis<br>Mild subluxation<br>L Thumb MCP |  |
|  |  |  |  |  |  |  | **Generalized ligamentous laxity** |  |

Details of individual MyCode participant phenotypes are presented in the supplemental materials (Variants associated with phenotype >40% of participants). Studies focused on this position of the gene and protein are warranted.

Of the other variants found to have phenotypes in greater than 40% of participants (Figure 4), none were reported in either the SVAS or TAAD series (Metcalfe et al.^18^, Overwater et al.^21^). Details of these variants including literature review and detailed phenotypic findings are presented in the supplemental materials (Variants associated with phenotype >40% of participants).

**Figure 4.**
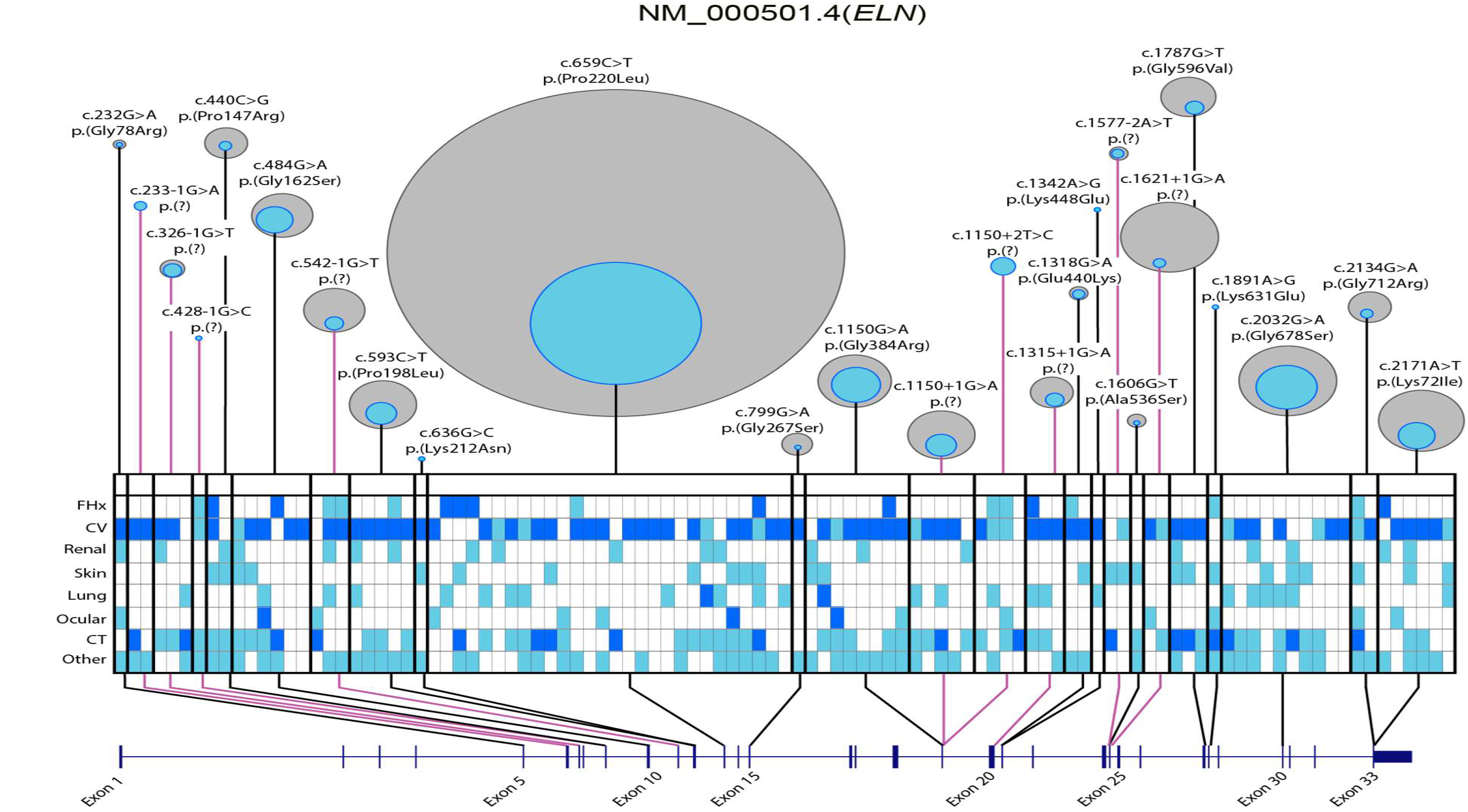
Variants with phenotype positive individuals and phenotype domains: (top) Gray circles represent total number of individuals with a variant listed in HGVS, inner blue circle represents individuals with phenotype (at least 1 major or three minor). Size of circles are proportional, and relative to each other across variants. Variants with no phenotype positive participants are not included (see Table S4 for full variant details). (bottom) Participants classified as having phenotype are shown in each column, each row is a phenotype domain. (FHx = family history, CV = cardiovascular, CT = connective tissue) In the phenotype grid, dark blue represents a major feature, cyan a minor feature, and grey absence of any feature. Variant positions are shown against the refseq NM_000150.4 transcript for ELN. Black connecting lines indicate missense, magenta indicate splice.

It is noteworthy that two of the missense variants with a higher proportion of phenotype-positive participants occupy the last nucleotide position of an exon. One of the missense variants reported by Metcalfe et al.^18^, NM_000501.4:c.55A>T also occupies the last nucleotide position of exon 3.

Splice variants were also noted in this study however Metcalfe did not report any of the splice variants seen in this study. This suggests aberrant splicing as a possible mechanism. Further studies of these variants to elucidate effects on splicing, protein structure and function, coupled with functional studies are needed.

### Insights from PheWAS

We identified two significant and robustly associated PheCodes in our PheWAS analysis, “arterial dissection” and “ganglion and cyst of the synovium, tendon, and bursa”. Aortic dissection, also noted by chart review, is a serious condition in which a tear occurs in the inner layer of the aorta, usually preceded by an aneurysm. Both ganglion cysts and bursae are cystic lesions found at the joints lined by a dense fibrous connective tissue and filled with viscous material^22,23^, distinguishable based on location (most commonly found in the wrist and knee, respectively) and presence of synovial cells in the case of bursae. These phenotypes (arterial dissection and cystic lesions) are similar in that stretching of weakened tissue is followed by rupture, herniation and/or inflammation^22-27^.

Additionally, both are associated with connective tissue abnormalities^28,29^. Abnormal joints including joint laxity and dislocation can be seen in people with pathogenic variants in *ELN* and ganglion cysts and bursae might fit into this spectrum^28^ further supporting the hypothesis that variation in *ELN* has connective tissue effects.

While no individual dermatologic trait reached a significance threshold in our analyses, the PheWAS was enriched for lower P-values in this category. Elastin is known to be involved in repair of cutaneous injury^30^ but susceptibility to skin infection has not been reported. Among our nominally associated dermatologic PheCodes was 686.1 “Carbuncle and furuncle” (OR=2.12, P=0.03). Carbuncles and furuncles are cutaneous infections that form bacterial abscesses.

Cutaneous infections are not a consistent feature in patients with haploinsufficiency in *ELN*^31^. We reviewed charts for this phenotype, but they were not consistently observed. However, unless severe and recurrent, they may not come to medical attention or be documented in medical records. Further investigation of these and other dermatological phenotypes may require more in-depth acquisition of medical history.

A GWAS for herniae^7^ identified significant associations with *ELN*, and other elastic fiber genes. This association was primarily driven by inguinal hernia. Based on this new discovery, and prior observation that about one-half of individuals with Williams-Beuren syndrome have an inguinal hernia^32^, the hernia PheCode and inguinal hernia diagnostic code were reviewed from our PheWAS. Neither were found to be significant (550 “abdominal hernia”, P=0.24; 550.1 “inguinal hernia, P=0.68). Specific information on the single nucleotide polymorphisms (SNPs) proximate to *ELN* that were driving the association was not provided, and it is possible that our ES analysis in MyCode would not have representation in this association region. While it is plausible that elastic tissue abnormalities could contribute to hernias involving the abdominal wall^33^, additional study is needed.

### Impact and Limitations

This study has some limitations. The chart review did not include controls without a variant in *ELN*. This decision was made for pragmatic reasons given the extensive nature of the chart review [Supplement File 1 Chart Review Form] and the attendant time required per participant. Inclusion of the PheWAS mitigates this weakness as does the use of population prevalence data for arterial dissection. The use of Z-scores to allow comparison of aortic diameters to population norms for an important phenotypic feature also addresses this concern.

Another significant weakness is related to diversity. Geisinger patients have a predominant genetic similarity to populations of Europe, reflecting historic patterns of settlement and low rates of in-and-out migration. While representing a population traditionally under-represented in research, (rural with lower socio-economic indices), it is not representative of the genetic, racial, and ethnic diversity of the United States as a whole. Extension of this research to more diverse populations is needed to ascertain the phenotypic impact associated with variation across race/ethnic and other genetic similarity groups.

Even though this is the largest group studied to date, the absolute number of variant carriers is still small. Thus, the PheWAS is underpowered to identify significant, but less prevalent phenotypes of interest.

The study relied on EHR data and illustrates both the strengths and weaknesses of these data for studies of this type. Compared to other cohorts, MyCode participants have both inpatient and outpatient records, imaging reports, and more extensive longitudinal data (mean of 14 years)^9^. However, the information recorded by clinicians is variable with much recorded as unstructured data in free text. Even when quantitative measurements are available, clinician interpretation may be biased. Not a single participant had documentation of hypoplasia of the aortic root or ascending aorta, despite this feature being present (by Z-score) in more than a quarter of participants that had an echocardiogram. This also impacts the PheWAS which relies on coded data and requires a minimum of two recorded instances of a billing code. Manual chart review somewhat mitigates this weakness, as it does include review of free text, but this analysis is qualitative thus may not improve the quality of the PheWAS. Additionally, milder phenotypes such as carbuncle/furuncle or other skin infections and pes planus may not be recorded in the EHR as problems if they are not causing significant symptoms.

Lastly age--while age at review was captured, phenotype development over time was not assessed as a variable in the chart review. Certain phenotypic features are known to increase with age (e.g., vessel tortuosity, valve incompetence, and cutaneous laxity), however there aren’t population standards that can be used as an analytic comparator. This is less likely to impact the PheWAS analysis as all analyses were adjusted for age.

In summary, this study reports the phenotypic associations of variation in *ELN* in a large, unselected population examined by two complementary approaches, chart review and PheWAS. It confirms the importance of elastin as a structural component of blood vessels, identifies a novel phenotypic association with ganglion and bursae cysts, and additional traits that warrant further investigation, including dermatologic traits such as carbuncle/furuncle. Further studies are needed to increase the size and diversity of study populations. Deep phenotyping of variant carriers, expansion of variants included in analysis, case-control studies of specific phenotypes, longitudinal assessment of phenotype development, and quantifying the effects of the splicing variants are also warranted.

### IRB Approval

All participants provided informed consent the MyCode Community Health Initiative as approved by the Geisinger Institutional Review Board^22,24^.

The elastin project described in this paper was reviewed and determined to be exempt by the Geisinger Institutional Review Board (IRB#: 2019-1139).

## Data Availability

All data produced in the present study are available upon reasonable request to the authors

## Acknowledgements

The authors would like to acknowledge the invaluable support and assistance of Heather Ramey in managing this project. Dr. Nephi Walton provided substantial input for the grant proposal and structure of the project. We thank all the participants of the MyCode Community Health Initiative Study (MyCode) and the MyCode Research Team. We thank the members of the Geisinger-Regeneron DiscovEHR Collaboration who have been critical in the generation of the genetic data used in this study.

## Author Contributions

MSW is the principal investigator and BAK is the site principal investigator of the project and both are responsible for funding. All authors reviewed and had final approval of the submitted manuscript and revision. MSW, BAK, AJ were responsible for study conceptualization. GB, JG, RZ, TK, and MSW performed the chart review. AJ and NJ performed the PheWAS analysis. MAK was the primary variant scientist and, in conjunction with BAK, TRL, AJ, and NJ, performed the variant annotation, prioritization, and review. TRL and AJ led data visualization. All authors meet the ICMJE Authorship Guidelines.

## Data and Code Availability

There are restrictions to the sharing of DiscovEHR genetic datasets related to the sharing agreement between Geisinger and the Regeneron Genetics Center. Chart review data contain individually identifiable information and can only be shared in aggregate. Most results are included in the manuscript and supplemental tables, but specific inquiries related to the chart review of for PheWAS full summary statistics can be directed to the corresponding author. All variants are included in the main body of the paper and supplemental tables. Variants for all living participants with appropriate consent have been confirmed in a clinical laboratory and deposited in ClinVar by that clinical laboratory.

## Web Resources

Online Mendelian Inheritance in Man #185500 Supravalvar Aortic Stenosis https://www.omim.org/entry/185500

Online Mendelian Inheritance in Man #123700 Cutis Laxa https://www.omim.org/entry/123700

MyCode ScoreCard https://www.geisinger.org/precision-health/mycode

weCall v1.1.2 https://github.com/Genomicsplc/wecall

National Marfan Foundation Aortic Root Z-Scores for Adults https://marfan.org/dx/z-score-adults/

Aorta Calculator from the Yale School of Medicine https://medicine.yale.edu/surgery/cardio/research/aorta_calculator_v6_by_tt_370821_4431_v2.xlsx

Online Mendelian Inheritance in Man #250215 Metaphyseal Acroscyphodysplasia https://www.omim.org/entry/250215?search=acroscyphodysplasia&highlight=acroscyphodysplasia

## References

1. Kozel, B.A., Mecham, R.P. (2019) Elastic fiber ultrastructure and assembly. Matrix Biol. 84, 31–40.

2. Cho, M.H., Ciulla, D.M., Klanderman, B.J., Hersh, C.P., Litonjua, A.A., Sparrow, D., Raby, B.A. and Silverman, E.K. (2009) Analysis of exonic elastin variants in severe, early-onset chronic obstructive pulmonary disease. Am. J. Respir. Cell Mol. Biol. 40, 751–5. 10.1165/rcmb.2008-0340OC.

3. Yang, S., Wang, T., You, C., Liu, W., Zhao, K., Sun, H., Mao, B., Li, X., Xiao, A., Mao, X. and Zhang, H. (2013) Association of polymorphisms in the elastin gene with sporadic ruptured intracranial aneurysms and unruptured intracranial aneurysms in Chinese patients. Int. J. Neurosci. 123, 454–8. 10.3109/00207454.2013.763803.

4. Ruigrok, Y.M., Seitz, U., Wolterink, S., Rinkel, G.J., Wijmenga, C. and Urbán, Z. (2004) Association of polymorphisms and haplotypes in the elastin gene in Dutch patients with sporadic aneurysmal subarachnoid hemorrhage. Stroke 35, 2064–8. 10.1161/01.STR.0000139380.50649.5c

5. Benjamins, J.W., Yeung, M.W., van de Vegte, Y.J., Said, M.A., van der Linden, T., Ties, D., Juarez-Orozco, L.E., Verweij, N., van der Harst, P. (2022) Genomic insights in ascending aortic size and distensibility. EBioMedicine. 75, 103783.

6. Deng, L., Huang, R., Chen, Z., Wu, L. and Xu, D.L. (2009) A study on polymorphisms of elastin gene in Chinese Han patients with isolated systolic hypertension. Am. J. Hypertens. 22, 656–62 10.1038/ajh.2009.53. 10.1038/ajh.2009.53.

7. Fadista, J., Skotte, L., Karjalainen, J., Abner, E., Sørensen, E., Ullum, H., Werge, T.; iPSYCH Group; Esko, T., Milani, L., et al. (2022) Comprehensive genome-wide association study of different forms of hernia identifies more than 80 associated loci. Nat. Commun. 13, 3200. 10.1038/s41467-022-30921-4.

8. Williams, M.S. (2022) Population Genetic Screening in Health Systems. Annu. Rev. Genomics Hum. Genet. 23, 549–567. 10.1146/annurev-genom-111221-115239.

9. Carey, D.J., Fetterolf, S.N., Davis, F.D., Faucett, W.A., Kirchner, H.L., Mirshahi, U., Murray, M.F., Smelser, D.T., Gerhard, G.S. and Ledbetter, D.H. (2016) The Geisinger MyCode community health initiative: an electronic health record-linked biobank for precision medicine research. Genet. Med. 18, 906–13. 10.1038/gim.2015.187.

10. Buchanan, A.H., Kirchner, L.H., Schwartz, M.L.B., Kelly, M.A., Schmidlen, T., Jones, L.K., Hallquist, M.L.G., Rocha, H., Betts, M., Schwiter, R., et al. (2020) Clinical outcomes of a genomic screening program for actionable genetic conditions. Genet. Med. 22, 1874–1882. 10.1038/s41436-020-0876-4

11. Dewey, F.E., Murray, M.F., Overton, J.D., Habegger, L., Leader, J.B., Fetterolf, S.N., O’Dushlaine, C., Van Hout, C.V., Staples, J., Gonzaga-Jauregui, C., et al. (2016) Distribution and clinical impact of functional variants in 50,726 whole-exome sequences from the DiscovEHR study. Science 2016 354, aaf6814. 10.1126/science.aaf6814.

12. Staples, J., Maxwell, E.K., Gosalia, N., Gonzaga-Jauregui, C., Snyder, C., Hawes, A., Penn, J., Ulloa, R., Bai, X., Lopez, A.E., et al. (2018) Profiling and Leveraging Relatedness in a Precision Medicine Cohort of 92,455 Exomes. Am. J. Hum. Genet. 102, 874–889. 10.1016/j.ajhg.2018.03.012.

13. Li, H. and Durbin, R. (2009) Fast and accurate short read alignment with Burrows-Wheeler transform. Bioinformatics 25, 1754–60. 10.1093/bioinformatics/btp324.

14. Yun, T., Li, H., Chang, P.C., Lin, M.F., Carroll, A. and McLean, C.Y. (2021) Accurate, scalable cohort variant calls using DeepVariant and GLnexus. Bioinformatics 36, 5582–5589. 10.1093/bioinformatics/btaa1081.

15. Staples, J., Qiao, D., Cho, M.H., Silverman, E.K.; University of Washington Center for Mendelian Genomics; Nickerson, D.A. and Below, J.E. (2014) PRIMUS: rapid reconstruction of pedigrees from genome-wide estimates of identity by descent. Am. J. Hum. Genet. 95, 553–64. 10.1016/j.ajhg.2014.10.005.

16. Wei, W.Q., Bastarache, L., Carroll, R.J., Marlo, J.E., Osterman, T.J., Gamazon, E.R., Cox, N.J., Roden, D.M. and Denny, J.C. (2017) Evaluating PheCodes, clinical classification software, and ICD-9-CM codes for phenome-wide association studies in the electronic health record. PLoS One 12:e0175508. 10.1371/journal.pone.0175508.

17. Wu, P., Gifford, A., Meng, X., Li, X., Campbell, H., Varley, T., Zhao, J., Carroll, R., Bastarache, L., Denny, J.C., Theodoratou, E. and Wei, W.Q. (2019) Developing and Evaluating Mappings of ICD-10 and ICD-10-CM Codes to PheCodes. JMIR Med. Inform. Sep 24. 10.2196/14325.

18. Metcalfe, K., Rucka, A.K., Smoot, L., Hofstadler, G., Tuzler, G., McKeown, P., Siu, V., Rauch, A., Dean, J., Dennis, N., et al. (2000) Elastin: mutational spectrum in supravalvular aortic stenosis. Eur. J. Hum. Genet. 8, 955–63. 10.1038/sj.ejhg.5200564.

19. Stamm C., Friehs I., Ho S.Y., Moran A.M., Jonas R.A., del Nido P.J. (2001) Congenital supravalvar aortic stenosis: a simple lesion? Eur. J. Cardiothorac. Surg. 19, 195–202. 10.1016/s1010-7940(00)00647-3.

20. Harkness, M. L., Harkness, R. D. and McDonald, D. A. (1957) The collagen and elastin content of the arterial wall in the dog. Proc. R. Soc. Lond. B Biol. Sci. 146, 541–551. 10.1098/rspb.1957.0029.

21. Overwater, E., Marsili, L., Baars, M.J.H., Baas, A.F., van de Beek, I., Dulfer, E., van Hagen, J.M., Hilhorst-Hofstee, Y., Kempers, M., Krapels, I.P., et al. (2018) Results of next-generation sequencing gene panel diagnostics including copy-number variation analysis in 810 patients suspected of heritable thoracic aortic disorders. Hum. Mutat. 2018 39, 1173–1192. 10.1002/humu.23565.

22. Gregush, R.E. and Habusta, S.F. 2023 Ganglion Cyst. In: StatPearls [Internet] Treasure Island (FL): StatPearls Publishing; 2023.

23. Jain, M., Sahu, N.K., Behera, S., Rana, R. and Patra, S.K. (2019) Intra-tendinous Patellar Ganglion Cyst Maybe the Unusual Cause of Knee Pain: A Case Report. Cureus 11:e5467. 10.7759/cureus.5467.

24. Telischak, N.A., Wu, J.S. and Eisenberg, R.L. (2014) Cysts and cystic-appearing lesions of the knee: A pictorial essay. Indian J. Radiol. Imaging 24, 182–91. 10.4103/0971-3026.134413.

25. Micheli, S., Paciaroni, M., Corea, F., Agnelli, G., Zampolini, M. and Caso V. (2010) Cervical artery dissection: emerging risk factors. Open Neurol. J. 2010 4:50–5. 10.2174/1874205X01004010050.

26. Haneline, M.T. and Rosner, A.L. (2007) The etiology of cervical artery dissection. J. Chiropr. Med. 6, 110–20. 10.1016/j.jcme.2007.04.007.

27. Blum, C.A. and Yaghi, S. (2015) Cervical Artery Dissection: A Review of the Epidemiology, Pathophysiology, Treatment, and Outcome. Arch. Neurosci. 2:e26670. 10.5812/archneurosci.26670.

28. McKeon, K.E., London, D.A., Osei, D.A., Gelberman, R.H., Goldfarb, C.A., Boyer, M.I. and Calfee, R.P. (2013) Ligamentous hyperlaxity and dorsal wrist ganglions. J. Hand Surg. Am. 38, 2138–43. 10.1016/j.jhsa.2013.08.109.

29. Erhart, P., Körfer, D., Dihlmann, S., Qiao, J.L., Hausser, I., Ringleb, P., Männer, J., Dikow, N., Schaaf, C.P., Grond-Ginsbach, C. and Böckler, D. (2022) Multiple Arterial Dissections and Connective Tissue Abnormalities. J. Clin. Med. 11, 3264. 10.3390/jcm11123264.

30. Baumann, L., Bernstein, E.F., Weiss, A.S., Bates, D., Humphreyc, S., Silberberg, M. and Daniels, R. (2021) Clinical Relevance of Elastin in the Structure and Function of Skin. Aesthet. Surg. J. Open Forum 3, ojab019. 10.1093/asjof/ojab019.

31. Kozel, B.A., Bayliss, S.J., Berk, D.R., Waxler, J.L., Knutsen, R.H., Danback, J.R. and Pober, B.R. (2014) Skin findings in Williams syndrome. Am. J. Med. Genet. A. 164A, 2217–25. 10.1002/ajmg.a.36628.

32. Li, F.F., Chen, W.J., Yao, D., Xu, L., Shen, J.Y., Zeng, Y., Shi, Z., Ye, X.W., Kang, D.H., Xu, B., et al. (2022) Clinical phenotypes study of 231 children with Williams syndrome in China: A single-center retrospective study. Mol. Genet. Genomic Med. 10, e2069. 10.1002/mgg3.2069.

33. Mosanya, A.O., Olasehinde, O., Odujoko, O.O., Etonyeaku, A.C., Adumah, C.C. and Agbakwuru, E.A. (2020) Comparative study of collagen and elastin content of abdominal wall fascia in inguinal hernia and non-hernia patients in an African population. Hernia 2020 24, 1337–1344. 10.1007/s10029-020-02238-y.

34. Pulignani S., Vecoli C., Borghini A., Foffa I., Ait-Alì L., Andreassi M.G. Targeted Next-Generation Sequencing in Patients with Non-syndromic Congenital Heart Disease. (2018) Pediatr. Cardiol. 39, 682–689. doi: 10.1007/s00246-018-1806-y.

